# Emergency Care Capacity and Accessibility for Non-Communicable Diseases in Kenyan EDs: A Regional Analysis

**DOI:** 10.64898/2026.02.23.26345636

**Authors:** Christine Ngaruiya, Guangyu Tong, Hani Mowafi, Lauren Hartz, Bhitariyo Mulimba, Mitali Shah, Jessica Rayo, Ivy Nyayieka, Benjamin Wachira

## Abstract

**Background:** Emergency Departments (EDs) are crucial to managing non-communicable diseases (NCDs), a leading cause of death, yet there is limited information about the capacity and accessibility of emergency care in Kenya.

**Methods:** Using data from Project 47, a national dataset on EDs, we conducted a secondary analysis to assess capacity for NCD care, as guided by the WHO Package for Essential NCD Interventions for Primary Care (WHO PEN).

**Results:** Of the 186 facilities included in the assessment, 45.7% (n=85) had a designated ED. ED capacity, distribution of facilities, number of patients seeking care, and WHO PEN indicators varied by Regional Economic Bloc (REB). Mount Kenya and Aberdares (MKAREB) and Lake Region (LREB) REBs included the greatest number of facilities and served the largest patient catchment areas nationally (8.5 million and 17.8 million, respectively). Key diagnostic tools and treatments were inconsistently available, with ECGs lacking outside of Nairobi (100% of EDs in Nairobi, 14.8% to 33.3% outside of Nairobi, P=0.1021). Oxygen was consistently available in only 44.8% of facilities in FCDC-Pwani, 59.3% in LREB, and 42.9% in NAKAEB, highlighting significant regional gaps, despite the LREB serving a disproportionately larger patient population. Glucometers, similarly, were universally available (100%) in Nairobi, but only available variably in other regional blocs (51.7% to 93.3%, P=0.001474).

**Conclusion:** Major disparities exist in emergency care capacity and NCD management resources across Kenyan regions, especially in LREB and NAKAEB, where majority of care occurs in the country. Targeted, equitable investment is required to expand essential infrastructure and diagnostics. Decentralizing emergency services beyond Nairobi must be prioritized to improve timely access to care, enhance quality, reduce patient costs, and address the needs of high-burden regions.

**KEY MESSAGES:** *What is already known:* Emergency departments in Kenya provide a key role in treating and managing non-communicable diseases. The most comprehensive data available about emergency department infrastructure, resources, and clinical visits in the country were collected via Project 47, conducted by the Emergency Medicine Kenya Foundation.

*What this study adds:* This secondary analysis of Project 47 data provides information about the capacity and accessibility of emergency care in Kenya, particularly as it pertains to Noncommunicable Diseases, focusing on cardiovascular disease, chronic respiratory disease, diabetes, and cancer, acute and emergency care, and the variability across different regions.

*How this study might affect research, practice, or policy:* Resources to improve the capacity and accessibility of emergency care in Kenya can be directed to the areas of highest need in order to maximize the impact on patient care. Similar assessments can be conducted in the region to guide emergency care resource allocation.

## INTRODUCTION

Deaths due to non-communicable diseases (NCDs) have surpassed those due to communicable diseases globally and are projected to do so in Africa by 2030.[1,2] Of the 40 million annual global deaths due to NCDs, 75% of these occur in low- and middle-income countries (LMICs).[1] Current trends predict worsening of the situation, with 55 million deaths from NCDs projected annually by 2030. Similarly, while NCDs currently account for 40% of deaths in East Africa, they are poised to become the leading cause of death by 2030.[2] The need to address NCDs in this region is critical.

Emergency Departments (EDs) in LMICs play a key role in the diagnosis and treatment of NCDs. This includes the acute stabilization of critically ill patients, as well as the initiation and adjustment of crucial medications like aspirin or antihypertensives, and the timely referral and linkage to care. Despite this pivotal role, there is a lack of data on current capacity for NCD care in EDs in LMICs. In 2018, the Emergency Medicine Kenya Foundation conducted Project 47, a cross-sectional study of all EDs in the 47 counties of the country, using two validated tools: the WHO Generic Essential Emergency Equipment List and the WHO Integrated Management for Emergency & Essential Surgical Care (IMEESC) toolkit.[3-5]

The objective of our study was to conduct a secondary analysis using results from the Project 47 study to assess capacity for and accessibility of NCD care in Kenyan EDs. The assessment of NCD care capacity was guided by the WHO Package for Essential NCD interventions for primary care (WHO PEN)[6], which highlights core components of care for the four lead NCDs in the primary care setting: cardiovascular disease, cancer, diabetes, and chronic respiratory disease. We supplemented the WHO PEN package with additional indicators that were available in the Project 47 dataset that we felt were essential for acute and emergent NCD care (e.g., stethoscope, blood pressure cuff, electrocardiogram (ECG), glucometer, and supplemental oxygen) to identify gaps in NCD care capacity by region across Kenyan EDs.

This is the first study of its kind in Africa, with potential outputs driving clinical guideline, intervention, and policy development to improve access to acute and emergent NCD care in Kenya. It also provides novel primary data to support development of NCD care across Africa and in emergency care settings in other low-resource settings globally.

## METHODS

### Setting

The setting of our study was in Kenya, which is a lower-middle-income country in East Africa with a population of 54 million people as of 2023.[7]

#### Kenya Regional Economic Blocs (REBs) & healthcare service delivery

The country is divided into 47 counties, and the capital city is Nairobi (Figure 1).[8] In turn, these 47 counties are grouped into seven “Regional Economic Blocs” (REBs). The REBs consist of counties that are geographically clustered together and that have similar cultural practices, language, and epidemiology profile that were grouped to increase political and economic capacity of individual counties. Furthermore, since health policy decisions occur at the level of the REB, we selected this as our unit of regional analysis for this study. The seven REBs are the Frontier Counties Development Council (FCDC), North Rift Economic Bloc (NOREB), Lake Region Economic Bloc (LREB), Jumuia ya Kaunti za Pwani (JKP), South Eastern Kenya Economic Bloc (SEKEB), Mt Kenya and Aberdares Region Economic Bloc (MKAREB), and the Narok-Kajiado Regional Economic Bloc (NAKAEB).[9] FCDC and JKP were collapsed for analysis due to overlap of sub-counties within these blocs.

**FIG 1:**
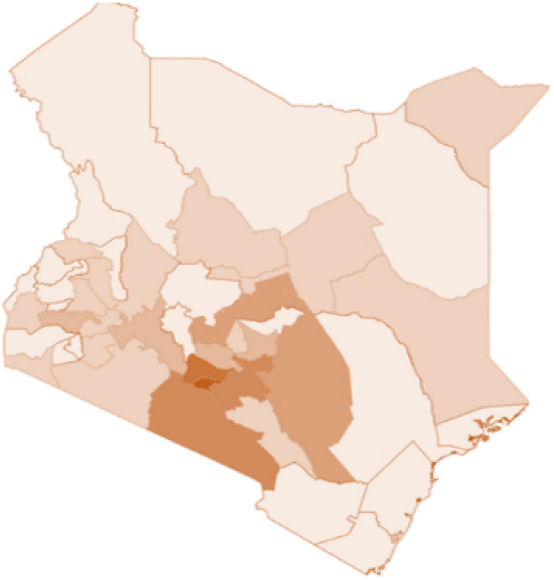
47 Kenya Counties

### Data Source: Project 47

Data for this analysis originated from the “Project 47” study. The methodology of that study is described in detail elsewhere.[3] Notably, the tools used in that study allowed for a broad assessment of emergency healthcare capabilities without delineation on specific disease types. Specifically, it assessed essential emergency equipment for resuscitation and minimum requirements for emergency and essential surgical care at health facilities (WHO Generic Essential Emergency Equipment List [2]), and gaps in availability of infrastructure and resources (WHO Integrated Management for Emergency and Essential Surgical Care (IMEESC) toolkit [5]). The existing Project 47 data was effective in our secondary analysis targeting NCD care specifically, as it provided key data on healthcare resources and emergency department capacity.

### Secondary analysis of Project 47 data

#### Inclusion criteria

Facilities were included in the secondary analysis if they a) had an ED as defined by Project 47 [3], or b) if those without a designated ED had the capacity to provide the equivalent level of care. For example, a facility may care for emergency patients in a unit equivalent to one of the facility’s outpatient clinics located adjacent to the hospital.

#### Facility type & resources

As the aim of the study was to assess access to services by the general public, we included only facilities that provide services in the public health system. All public level 4, 5, and 6 facilities in the dataset were included; these constitute sub-county, county, and national referral hospitals, respectively. Mission hospitals and non-governmental organization (NGO) hospitals were also included, as emergency care in these facilities is provided for free or reduced fees equivalent to those of the public health system. Privately owned healthcare facilities were excluded due to their high costs and limited geographic coverage, which limited their access for the general public.[14,15]

#### ED capacity

We assessed ED capacity based on designated space for care and availability of trained providers. This was indicated by the distribution of ED beds and ED physicians, specifically. The availability of resources for NCD care in emergency departments (EDs) was assessed using the WHO PEN checklist by mapping its components onto corresponding items in the original dataset.[6]

#### Patient characteristics

We conducted a descriptive analysis of patient access to facilities. This included the number, gender, and age of patients seeking care at included facilities. We further classified patients seeking care by Regional Economic Bloc, level of healthcare facility (levels 4-6 as described above), type of healthcare facility (public, charity/mission, or NGO as described above), and the associated care capacity at those facilities (indicated by beds per capita). Finally, we described the breakdown of patients by disposition: the proportion of outpatients, admissions, and referrals. This informs the level of acuity or severity of illness in EDs. Chisquared tests with Yate’s continuity correction were employed to assess disparities across regions. The analysis was conducted using R 4.2.1.

#### Variable creation

The regional average was created by calculating the average of facility-level statistics within each region. Instances of missing data were excluded from the averaging. Of note, FCDC and JKP were collapsed into one unit for analysis, given overlap of two sub-counties in their respective REBs (i.e., Lamu County and Tana River). Nairobi was isolated for analysis given its uniqueness. The capital city consists of a predominantly urban population and is generally disproportionately higher resourced when compared to the remainder of the country.[12] Only two facilities from Nairobi met inclusion criteria for our study, yet the proportion of care that occurred in those two hospitals alone was high, with the number of admissions in Nairobi alone matching that of an entire REB (NAKAEB), see Table 2.

## RESULTS

Demographic information for the seven REBs and Nairobi are presented in Table 1. Kenya has six levels of healthcare facilities that range from those that provide community-based services (Level 1) to those that provide services at the national level (Level 6).[10] Only Level 4, 5, or 6 facilities in Kenya would have the capacity to resuscitate, diagnose, and treat patients with medical emergencies and meet the WHO definition of an ED (any dedicated intake area for acutely ill and injured patients).[4] According to the Kenya Ministry of Health, there are currently 518 public and private Level 4, 5, and 6 facilities across the country [10].

**Table 1:** Demoratic and healthcare capacity information for regional analysis units (Kenya Regional Enocomics Blocs and Nairobi)

|  | FCDC & JKPa | LREB | MKAREB | Nairobi | NOREB | SEKEB | NAKAEB |
| --- | --- | --- | --- | --- | --- | --- | --- |
| Total population[11] | 7,547,334 | 14,944,940 | 11,103,668 | 4,397,073 | 6,019,025 | 3,545,772 | 2,275,713 |
| Gross County Product, per capita (USD)[12] | 1,381 | 1,497 | 2,179 | 6,182 | 1,456 | 1,444 | 1,390 |
| Purchasing Power Parity, per capita (USD) [13] | 3,761 | 4,077 | 5,934 | 16,837 | 5,287 | 4,353 | 3,786 |
| Number of hospitals, by level* |  |  |  |  |  |  |  |
| Level 3 | 4 | 7 | 2 | 0 | 2 | 0 | 0 |
| Level 4 | 18 | 31 | 41 | 2 | 15 | 13 | 4 |
| Level 5 | 7 | 13 | 13 | 0 | 4 | 2 | 3 |
| Level 6 | 0 | 3 | 2 | 0 | 0 | 0 | 0 |
| Number of hospitals, by type* |  |  |  |  |  |  |  |
| Public | 27 | 38 | 36 | 2 | 13 | 13 | 5 |
| Mission | 0 | 15 | 22 | 0 | 8 | 2 | 1 |
| NGO | 2 | 2 | 0 | 0 | 0 | 0 | 1 |
| Number of hospital beds (bed capacity)* | 3,028 | 7,932 | 8,797 | 360 | 2,166 | 1,975 | 547 |
| Number of ED beds (bed capacity)* | 86 | 156 | 163 | 16 | 45 | 37 | 10 |

\*FCDC and JKP collapsed for analysis due to overlap of sub-counties within these blocs \*As presented in the Project 47 database; not reflective of total number of country hospitals, distribution/ capacity. Table: Stanford Department of Emergency Medicine • Source: Project 47 • Created with Datawrapper

**Table 2:** Patient demographics and care capacity, by REB*.

|  | FCDC + JKP | LREB | MKAREB | Nairobi | NOREB | SEKEB | NAKAEB |
| --- | --- | --- | --- | --- | --- | --- | --- |
| Total population served by REB | 1,728,927 | 17,874,960 | 8,501,372 | 395,519 | 587,636 | 2,128,155 | 1,520,014 |
| Male | 586,137 | 2,764,805 | 3,362,992 | 205,670 | 241,201 | 1,157,586 | 137,362 |
| Female | 541,888 | 3,296,808 | 3,738,164 | 189,849 | 281,647 | 1,110,616 | 74,652 |
| Adult admissions per year | 73,073 | 491,126 | 2,388,768 | 25,831 | 142,134 | 41,058 | 25,531 |
| Adult patients seen in ED per year | 91,192 | 312,045 | 268,990 | 2,200 | 25,343 | 14,740 | 2,958 |
| Percent adult ED patients referred to higher-level facility per year | 1.78% | 1.83% | 2.89% | 3.95% | 5.88% | 8.19% | 8.55% |
| Child (<15 years) admissions per year | 26,534 | 89,154 | 985,054 | 6,315 | 78,014 | 10,665 | 5,972 |
| Child (<15 years) patients seen in ED per year | 14,805 | 32,150 | 33,669 | Not Available | 6,412 | 4,224 | Not Available |
| Percent child (<15 years) ED patients referred to higher-level facility per year | 5.03% | 7.83% | 6.29% | Not Available | 18.20% | 12.20% | Not Available |
| Number of ED physicians | 139 | 259 | 332 | 73 | 43 | 61 | 25 |
| Number of ED Clinical Officers | 202 | 455 | 591 | 60 | 92 | 107 | 29 |

aFCDC and JKP collapsed for analysis due to overlap of sub-counties within these blocs
\*As presented in the Project 47 database; not reflective of total number of country hospitals, distribution/ capacity.
Table: Stanford Department of Emergency Medicine • Source: Project 47 • Created with Datawrapper

Of the 518 level 4, 5, and 6 facilities in Kenya, a total of 186 facilities met inclusion criteria for this study. Of those facilities meeting inclusion criteria, there was a 98.5% response rate in the Project 47 study. Overall, of the 186 facilities included in the assessment, less than half (45.7%, n=85/186) had a designated ED. Of all the REBs, MKAREB had the highest proportion of facilities with a designated ED (56.9% of facilities in that REB, n=33/58). This proportion was exceeded only by Nairobi, where both included facilities had a designated ED. MKAREB also had the overall highest number of facilities with a designated ED as compared to all other regions (n=33), representing more than a third of all facilities with an ED nationally in our dataset.

There were five level-6-facilities (national referral hospitals) included. These were located in MKAREB (n=2), which also had the highest number of overall facilities, and in the LREB (n=3). The majority of included facilities were level 4 (sub-county/ acute general hospital; n=124) or level 5 (county referral hospital; n=42) facilities.

The majority of facilities in our study were public hospitals (n=134; Table 1). The MKAREB had the largest number of facilities included overall (n=58), 36 of which were public. LREB had the second highest number of facilities (n=54), and nearly a third of hospitals were run by non-governmental entities in this region. NAKAEB, which is also the smallest bloc consisting of only two counties, had the least number of healthcare facilities (n=7); two of the seven facilities were non-public—one mission hospital and one NGO hospital. Nairobi city had only two facilities in the dataset, both of which were public hospitals.

There were a total of 32,736,583 people in the catchment area of the 186 facilities (Table 2). While gender data was not recorded for almost half of the catchment population, women were a slight majority of those recorded. Among those actually reported to have sought care, just 2.19% of adults and 0.28% of children received care in a designated ED. However, there was a high percentage of missing data on age (102 facilities missing adult data, 125 facilities missing pediatric data).

By region, the largest catchment areas were in LREB (n=17,874,960) and MKAREB (n=8,501,372, 77% of population). The smallest catchment areas were in the NOREB (n=587,636). 395,519 people constituted the catchment area at facilities in Nairobi alone, despite only two facilities meeting inclusion criteria in the capital city for this study. Of note, the number of hospital beds (total in all regions, n=24,805) were also highest in the MKAREB (n=8,797) and LREB (n=7,932) regions (Table 1). The mean patient-to-bed ratio was 1.3 patients per capita (s.d. +/-1.15). The highest ratio of beds per capita was in NOREB (3.7 beds per 1,000 patients, n=2166 total hospital beds/ 395,000 people being served in the county); the lowest ratios were in LREB with a ratio of 0.44 and NAKAEB with a ratio of 0.36 beds per 1,000 patients, respectively. Only 513 or 2.07% of all reported hospital beds nationally were reported to be ED beds (Table 1).

As far as acuity, there were a total of 3,187,500 patients admitted (13% of all patients seeking care, 8 counties missing); the most admissions occurred in MKAREB and LREB. Overall, 18,143 adult patients were referred to a higher-level of care facility from the ED (Table 2). The highest proportion of referrals occurred in NAKAEB and SEKEB, and the lowest from FCDC. SEKEB had the lowest proportion of designated ED beds of any bloc; NAKAEB and SEKEB also had the least number of ED physicians by region. NAKAEB had the highest proportion of level 5 facilities as compared to lower level facilities of any bloc, which indicates a high level of referral, even though they have these higher level sites that should be able to manage sick patients. Of note, the two blocs with level 6 facilities (LREB and MKAREB) had the lowest proportion of ED referrals.

Five key indicators were selected from the WHO PEN to assess NCD care capacity: presence of a stethoscope, blood pressure cuff, ECG, glucometer, and oxygen supply. We found statistically significant differences in the availability of core NCD diagnostics by region, except for ECG (see Table 3). Stethoscopes were least accessible in the LREB (always available only two-thirds of the time, or 64.8% of the time in the 35 facilities included in the analysis). They were reported to be universally available at facilities in Nairobi and NAKAEB. Blood pressure cuffs were least accessible in FCDC-Pwani (65.5% reported having BP cuffs always available), LREB (66.7%), and NOREB (76.2%). ECGs were universally poorly accessible (no statistically significant difference across all regions), reported in only around a quarter of EDs in all sites except Nairobi, where both facilities reported always having ECG available. Glucometers, similar to blood pressure cuffs, were less accessible in FCDC-Pwani and LREB, only around half of the time. Finally, while oxygen was also reported to be always available in the Nairobi facilities, it was available only around half of the time in FCDC-Pwani (44.8%) and LREB (59.3%), and less than half of the time (42.9%) in NAKAEB.

**Table 3:**
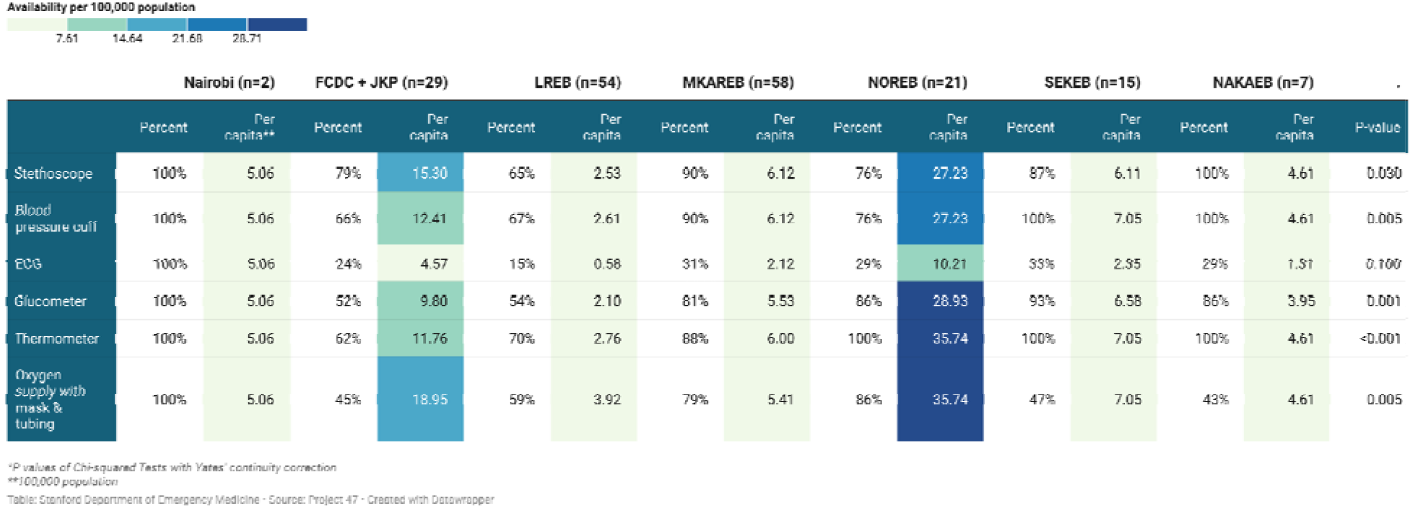
The availability of WHO PEN technologies by region.

## DISCUSSION

Overall, access to designated emergency departments is limited in Kenya. This is evidenced by the number of facilities with a separate designated area to care for emergency patients (“an emergency unit”). Furthermore, even among those facilities with an emergency unit, there is regional variation in the availability of ED-designated beds compared to total visit volume and area population, with some of the most populated areas having a very low availability of ED beds per patient volume. This causes ramifications for management and stabilization of acute and emergency conditions, spanning public health conditions of importance such as trauma and injury, as well as communicable and noncommunicable diseases; it can be reasonably inferred that patients who present in areas not designated as emergency units are likely to receive a lower standard of acute or emergency care.

The presence of a designated ED or emergency unit in LMICs is essential for response to high acuity patients and for improving patient outcomes.[16-18] Patients with AMI, for example, that receive care in a designated cardiac care center (including an equipped ED) have better outcomes [19], and similar results have been shown for other noncommunicable diseases.[20,21] In short, this supports the establishment of emergency departments that are appropriately resourced through staffing as well as diagnostic tests to ensure appropriate care for patients with the most emergent conditions to improve patient outcomes in terms of lower rates of morbidity and mortality from high acuity conditions, with implications for economic stability and population-level health improvement if this occurs [22].

The development of emergency care remains a standing call for other African countries also. In Rwanda, many gains have been made in various aspects of development, but there are still gaps in the delivery of emergency care.[23] Studies of the availability of emergency care in Tanzania have shown gains over the past decade [24-26], and low-cost interventions show promise in further promoting emergency care knowledge.[27] Small-scale studies in countries such as Eswatini[28] and Cameroon[29] have shown both strengths and challenges, but large-scale data collection and analyses are limited across Africa. Frameworks such as Project 47 and secondary analyses of data collected in these large-scale projects will be necessary for understanding the current state of emergency care in Africa, providing benchmarks for comparison, and designing improvement programs that effectively and efficiently meet the needs of individual countries and regions.

Within Kenya, the regional variation of care need and ED availability was concerning. Furthermore, this data is not widely available to the public, whereas it could facilitate patient navigation for ED care based on distance, types of resources (skills or diagnostic tests), or other factors. This is a nationwide problem that should be addressed. Specifically, the majority of care occurred in two of the regional economic blocs, the Mount Kenya and Aberdares Regional Economic Bloc (MKAREB) and the Lake Region Economic Bloc (LREB), respectively. These two regional economic blocs also constitute the largest proportion of the population nationally, with 11.1 million people residing in the MKAREB region and 14.9 million people residing in the LREB region.[11] However, the proportion of patients seeking care in these two regions differed. Of note, there was a disproportionately larger number of patients reported seeking care in the LREB region than in the MKAREB region. In addition, the LREB had a disproportionately lower patient-to-bed ratio when compared to MKAREB and other regional economic blocs, suggesting that the patients seeking care in this region may have limited access to admissions in their region, barriers to in-hospital care, and overall worse patient outcomes [30]. We propose that national efforts should include increasing standardization for ED capacity by region and increasing patient education on the capacity of care that may be available by region. Factors that can also be considered in assigning ED and bed capacity include demand for services, ability to access services, inter-regional patient flow, staffing availability, and availability of resources by hospital or county.

We propose that some of the variation observed in patient care access patterns, such as a disproportionately higher population seeking care in LREB over MKAREB may be related to perceived quality of care [31]. These care patterns may also be determined by the economic stability of the respective REB. For instance, the economy in the MKAREB bloc is reported as being more stable than the economy of LREB (see Table 1). This likely correlates with increased investment in healthcare facilities and emergency departments, which would contribute to an increased number of beds and higher patient-to-bed ratios, as was seen in our data (in MKAREB as compared to LREB). However, our data does not fully capture disproportionate patient patterns in access care, and factors driving these patterns should be further assessed, such as through community-based surveys and facility-level assessments. In addition, resource allocation should focus on those regions with a disproportionately higher patient population seeking care there.

Other regions also had low capacity for emergency patients (as defined by bed to patient ratio), such as NAKAEB. Therefore, these two regions in particular (LREB and NAKAEB) may need further infrastructure implicated in their county health budgets, as well as national allocation of resources. Further studies assessing patient preference or migratory patterns between regions or particular facilities may also provide key guidance on care-seeking behaviors that lead to targeted interventions addressing them. Common factors influencing care that may be intervened upon could include: improving quality of care across regions, better understanding care-seeking behaviors by disease or chief complaint type that may lead to improvements in capacity for certain types of care regionally, among other opportunities for improvement.

This analysis also confirmed that emergency care is highly concentrated in Kenya, as signalled by the relatively large proportion of patients being seen in the capital city, despite Nairobi only having two facilities included in this analysis, whereas several blocs had entire counties (multiple cities or towns included). This supports other evidence that Nairobi provides a leading proportion of clinical care in Kenya.[32,33] In the long term, decentralizing care will be key for timely access to care, including for care of NCDs such as acute myocardial infarction or strokes, in which time to care is of the utmost importance. The majority of facilities are still well-known to be outside of the 2-hour international benchmark for access to emergency care [34].

The vast majority of emergency care in Kenyan hospitals occurs in public facilities. However, there is a disproportionately high number of non-governmental organizations that provide care to populations in some of the most vulnerable areas with some of the poorest communities that otherwise have no options for access to care. While the obvious argument is to increase infrastructure for healthcare facilities by region more equitably, it is also important to ensure that the standard of care is being met at each type of facility, regardless of whether it is a non-governmental or other entity.[35,36] One of the difficulties with having nearly a third of non-governmental facilities constitute the source for care is that they may not be as regulated in terms of the type or quality of care being provided;[37] mission hospitals are subject to variations in healthcare personnel, the longevity with which they are on site in a given setting, and subsequent variations in clinical care that affects both short and long term effects on their patients. Some of the short-term effects include employing the use of variably trained individuals. In addition, there is the pervasive unintended consequence of varying from the standard of care within the country such as providing expensive procedures or tests that are otherwise not sustainable beyond that particular facility or set of individuals that may be available at a given time which then leaves the individuals who are receiving care shortchanged for continuity of initial care received at a mission hospital [38]. In sum, it will be imperative that the government ensures clinical guidelines for emergency care are rigorous, participatory to ensure sustainability, and widely enforced across facility type.

When describing acuity as indicated by admissions and referrals, we found that just 13% of patient visits result in admissions, which is in line with larger health systems like the US.[39,40] Admission rates in this context, naturally, are a poor marker for the severity of illness, given multiple additional factors that determine admission versus discharge outside of the illness or severity of illness at the time of evaluation. Some of these factors include the availability of resources to provide admission services, such as beds, and the ability of the patient or family to afford admission, which may be precluded at the time of the visit if funds are not available. However, this data point is still encouraging from the standpoint of improvements in capacity of care being attainable with the starting point being a relatively low proportion of patients warranting admission for visits in the country. Moreover, it was noted that there was a higher proportion of referrals among facilities and regions where they had fewer hospital beds, fewer designated emergency department beds, and fewer ED physicians. Based on this, and an implied assumption that the majority of care being provided is to lower acuity patients, measures to improve evaluation and care of lower-level acute/emergent patients should be prioritized as resources are being allocated. Options include interventions such as keeping outpatient clinics open later to take low acuity unscheduled visits instead of in the ED or ensuring appropriate messaging on the use of lower acuity facilities is communicated to the general population.

Finally, when considering key indicators for NCD care, it was alarming that one of the most populous counties (LREB) had the worst indicators of access to key care items such as stethoscopes, blood pressure cuffs, and glucometers – performing among the lowest across indicators when compared to other facilities. ECG access, critical to first-line diagnosis of multiple NCD conditions, including AMI, the leading cause for global mortality, had abysmal access in most facilities; this requires immediate national attention and action. ECGs should be implemented at all facilities, and associated training should be provided to ensure appropriate diagnosis and referral to care as needed. Oxygen supply was also dismal; this was possibly affected by the COVID-19 pandemic, but further resource allocation is needed to ensure acute oxygen access, such as for the care of acute asthma exacerbation, among other conditions. As expected, Nairobi outperformed most facilities for NCD diagnostic tool access, which further supports the argument on the need to decentralize acute and emergent NCD care, particularly as these are time-dependent conditions that require timely access to these core interventions.

## CONCLUSION

In total, this analysis provided an illustration of the care patterns for patients seeking emergency care in Kenya and revealed a need for strengthening the infrastructure and resources underlying this critical component of the country’s health system. This should be done in a way that is regionally equitable to ensure that unnecessary patient resources or costs are not spent on travel to neighboring regions, as is apparent in mismatched care by region in our dataset. This may include, as previously mentioned, ensuring appropriate staffing benchmarks (numbers and type of staffing, including but not limited to trained ER physicians, trained clinical officers, and nurses on acute/ emergency care), resources for care, especially for stabilization, and guidelines for care to standardize care occurring nationally. These interventions could especially be key for the penultimate level of facilities (level 5 hospitals) that constitute county level referral hospitals that are distributed across each of Kenya’s 47 counties (one per county) and generally centrally accessible for each county’s population, even if level 6 facilities are much less accessible by distance. Additional strategies should also include routine monitoring and evaluation, such as through registries on the distribution of patients seeking care, as well as types of diseases being addressed by facility and region.

## Data Availability

Data used in this study derive from Project 47 and are not publicly available. De-identified data may be made available upon reasonable request to the corresponding author, contingent on approval by the data custodians and any applicable ethics/data use requirements.

## ACKNOWLEDGEMENTS

We would like to acknowledge Mr. Mugane Mutua for his contributions during data collection, and Kimberly Korwek, PhD, for editing support.

